# Estimating the effectiveness of COVID-19 vaccination against COVID-19 hospitalisation and death: a cohort study based on the 2021 Census, England

**DOI:** 10.1101/2023.06.06.23290982

**Authors:** Charlotte Bermingham, Vahe Nafilyan, Nick Andrews, Owen Gethings

## Abstract

**Objective:** To estimate the effectiveness of COVID-19 vaccination against hospitalisation for COVID-19 and death involving COVID-19 in England using linked population level data sources including the 2021 Census.

**Design:** Retrospective cohort study.

**Setting:** England, 21 March 2021 to 20 March 2022.

**Participants:** Individuals alive and aged 16+ on 21 March 2021, resident in England, enumerated in the 2021 Census as a usual resident, and able to link to an NHS number. A sample of 583,840 individuals was used for the analysis.

**Exposures:** COVID-19 vaccination: first dose, second dose and third dose/first booster dose, with categories for time since each dose.

**Main outcome measures:** Hospitalisation for COVID-19 or death involving COVID-19. An adjusted Cox proportional hazard model was used to estimate the hazard ratio for the outcomes for vaccinated participants for different doses and time since dose compared to unvaccinated individuals. Vaccine effectiveness was estimated as (1–hazard ratio)×100%. A control outcome of non-COVID-19 death was also assessed.

**Results:** Vaccine effectiveness against hospitalisation for COVID-19 was 52.1% (95% confidence interval 51.3% to 52.8%) for a first dose, 55.6% (55.2% to 56.1%) for a second dose and 77.6% (77.3% to 78.0%) for a third dose, with a decrease in vaccine effectiveness 3+ months after the third dose.

Vaccine effectiveness against COVID-19 mortality was 58.7% (52.7% to 63.9%) for a first dose, 88.5% (87.5% to 89.5%) for a second dose and 93.2% (92.9% to 93.5%) for a third dose, with evidence of waning 3+ months after the second and third doses.

For the second dose, which is the most comparable across the different time-periods, vaccine effectiveness was higher against COVID-19 hospitalisation but slightly lower against COVID-19 mortality in the Omicron dominant period than the period before the Omicron variant became dominant. Vaccine effectiveness against both COVID-19 hospitalisation and mortality was higher in general for mRNA vaccines than non mRNA vaccines, however this could be influenced by the different populations given each vaccine vector. Non-zero VE against non-COVID-19 mortality indicates that residual confounding may impact the results, despite the inclusion of up-to-date socio-demographic adjustments and various sources of health data, with possible frailty bias, confounding by indication and a healthy vaccinee effect observed.

**Conclusions:** The vaccine effectiveness estimates show increased protection with number of doses and a high level of protection against both COVID-19 hospitalisation and mortality for the third/booster dose, as would be expected from previous research. However, despite the various sources of health data used to adjust the models, the estimates for different breakdowns and for non-COVID-19 mortality expose residual confounding by health status, which should be considered when interpreting estimates of vaccine effectiveness.

## Introduction

On the 8th December 2020, the United Kingdom’s Medicines and Healthcare products Regulatory Agency (MHRA) granted emergency use of the mRNA Covid-19 vaccine BNT162b2 (Pfizer-BioNTech) to “address significant public health issues” under the Medicines Act 1968 [1]. Further vaccines were approved shortly after including the ChAdOx1-S (Oxford/AstraZeneca) and mRNA-1273 (Moderna) becoming the second and third approved for use in the UK. Estimating real-world vaccine effectiveness (VE) is vital to assess the impact of the vaccination programme on the COVID-19 pandemic and inform the ongoing policy response. Large clinical trials have been undertaken for all of the vaccines approved in the UK, which found high efficacy against symptomatic disease and severe outcomes [2]–[4]. However, real-world post-implementation studies are necessary in order to understand vaccine effectiveness within different populations and against new and emerging variants.

Many estimates of vaccine effectiveness have been produced using large observational studies around the globe [5]–[9]. However, estimating vaccine effectiveness using observational data is challenging due to the non-randomised design and the potential for unmeasured confounding [10]–[12]. Many factors may be related both to the eligibility and decision to be vaccinated and the outcome of interest, such as age, ethnicity, occupation, and health status. If these factors are not adjusted for, it could lead to biases that can then either strengthen, weaken, or reverse the actual exposure-outcome relationship. To some extent, these effects can be accounted for by adjusting models for characteristics associated with likelihood to be vaccinated and the outcome of interest, but not all these factors are available in population-level data.

In this analysis we estimated vaccine effectiveness using a cohort study based on the 2021 Census linked to electronic health records, adjusting for a wide range of confounding factors. Using data from the 2021 Census provided up to date, population level data to adjust for confounding variables that can change for individuals over time, including socio-demographic factors such as English language proficiency and occupation or subjective measures of health such as self-reported health and disability. Therefore, the models can be better adjusted for possible confounding to produce more accurate estimates of VE.

## Methods

### Study data

We used a person-level dataset comprising of individuals in the 2021 Census, linked to the Personal Demographics Service [13] to obtain NHS numbers with a linkage rate of 91.1%. These individuals were then linked via NHS number to ONS death registrations [14] as well as to COVID-19 vaccination records, GP records, and hospital records. Vaccination records were obtained from the National Immunisation Management Service (NIMS) [15] and a supplementary extract from NHS-Digital point of care data of 1,069 new vaccinations of people in our linked dataset of 42 million people, where the person had died soon after vaccination and the vaccination was not included in NIMS.

Health related variables from primary care records were derived from the General Practice Extraction Service (GPES) data for Pandemic Planning and Research version 3 (GDPPR) and linked via NHS number [16]. The linked dataset included data on 52 million people resident in England, which covers approximately 91.7% of the population of England on Census day in 2021.

The Hospital Episode Statistics (HES) dataset for admitted patient care was used to obtain hospitalisation related variables [17]. [15]The Hospital Episode Statistics data covered episodes that ended up to 30 September 2022, the NIMS data covered vaccinations up to 22 February 2023 and the mortality data covered deaths registered up to 4 January 2023.

### Study population and time-period

The study population was individuals who were enumerated in the 2021 Census, were resident in England and alive and aged 16+ on the day of the 2021 Census (21 March 2021) and had an entry in the Personal Demographics Service. The linked dataset included data on 52 million people resident in England, which covers approximately 91.7% of the population of England on Census day in 2021. A sample size flow diagram is presented in **Supplementary Table 1**.

Individuals were followed from 21 March 2021 until the 20 March 2022, the date of the start of the spring booster campaign. As our dataset does not include spring booster doses, we would not be able to categorise vaccination status accurately beyond this date.

Data engineering and dataset construction was carried out using Python version 3.6.8, Spark version 2.4.0 and R version 3.5.1.

### Outcomes

The first outcome of time to COVID-19 hospitalisation was defined as an inpatient episode in HES where the primary diagnosis was COVID-19. The date of the start of the episode was used as the date of COVID-19 hospitalisation. Where an individual had experienced more than one COVID-19 hospitalisation, the earliest that occurred within the study period was used. The second outcome of time to death involving COVID-19 was defined as a death where the underlying or a contributory cause of death was recorded as COVID-19 on the death certificate. The date of death occurrence was used as the date of COVID-19 death. In both cases, COVID-19 was defined by the ICD-10 codes U07.1 or U07.2 and time at risk was the time from 21 March 2021 to the outcome occurring or end of study for individuals who did not experience the outcome. Finally, vaccine effectiveness against non-COVID-19 death was calculated as a negative control outcome to investigate the potential impact of residual confounding on the main results.

### Exposures

The exposure was COVID-19 vaccination status. The dose and the time since dose were accounted for using a time-varying variable for vaccination status. Participants moved group according to the dose received and contributed time at risk to the various groups. The ‘21 days to 3 months’ categories are referred to as first/second/third dose and later time periods after vaccination are included to see how the vaccine performs over time. The vaccine status groups are defined in **Supplementary Table 2**.

### Covariates

A range of sociodemographic and health related variables were used to adjust the model for factors likely to confound the relationship between vaccine status and COVID-19 hospitalisation or death (**Supplementary Table 2**). Vaccination uptake varies substantially by socio-demographic and economic characteristics, such as ethnicity, religious affiliation or occupation COVID-19 mortality has been shown to vary across these groups [18]–[21].

Socio-demographic and self-reported health variables were sourced from the 2021 Census. Fixed health-related variables used for model adjustment were BMI category, number of health conditions shown to impact the risk of COVID-19 hospitalisation and death in the QCovid risk model[22] and hospitalisation for a frailty related condition. We also included a time-varying covariate for whether an individual was currently in hospital or had been in hospital in the preceding 21 days to account for time varying health status. The variable is 1 on the day of hospital admission and switches to 0 21 days after discharge. The variable does not switch if there are overlapping periods of hospitalisation until 21 days after the final discharge that does not overlap with any other periods of hospitalisation.

### Statistical analyses

Characteristics of the baseline covariates for the study population were summarised, stratified by age group.

Hazard ratios for the outcomes were estimated using a Cox Proportional Hazard regression model adjusted for confounding factors, unless otherwise stated. The start of the time at risk was 21 March 2021. Vaccine effectiveness against COVID-19 hospitalisation and death was calculated as the complement of the hazard ratio (1-HR) times 100, with corresponding 95% confidence intervals.

Analyses were stratified by age group (16-29, 30-64, 65-79, 80+). Differences due to changes in the dominant COVID-19 variant in circulation were investigated by interacting vaccination status with a time varying variable for variant period, defined as pre-omicron (start of study 21 March to 19 December 2021) and omicron (20 December 2021 to end of study 20 March 2022). These results were also produced by variant and age group for the COVID-19 hospitalisation. The effect of vaccine vector was investigated using a time-varying covariate taking into account both vaccine vector and time since dose as the exposure.

For computational reasons, we used a sample of the full dataset to run the analyses. Individuals were weighted with a weighting inversely proportional to their probability of being sampled. The sample used for running the model included all who died from or were hospitalised for COVID-19 or were aged under 30 and died of a non-COVID-19 cause, a weighted 10% sample of those aged 30+ who died from a non-COVID-19 cause and were not hospitalised for COVID-19 and a weighted 1% sample of those who did not die and were not hospitalised for COVID-19.All analysis was carried out using R version 3.5.1. Cox proportional hazard models were carried out using the package ‘survival’ version 3.3.1.

### Sensitivity tests

We compared results from models with age only, age and socio-demographic characteristics and a fully adjusted model. The impact of including a time-varying covariate for current or recent hospitalisation on the analysis of COVID-19 hospitalisation was checked by omitting this variable from the analysis. The impact of including the first 21 days after a first dose of a vaccine in the ‘unvaccinated’ category was investigated by omitting these days from the ‘unvaccinated category’ (treating the 21 days after first dose as a separate vaccination status).

### Patient and public involvement

We did not directly involve patients and the public in the design and conception of the study, primarily because of the pace at which this study was conducted to inform the UK government’s response to the covid-19 pandemic. However, the paper was read by several members of the public.

## Results

### Characteristics of the study population

Our full individual level dataset contained 41,855,563 individuals, including 27,270 deaths involving COVID-19, 433,670 non-COVID-deaths and 115,495 COVID-19 hospitalisations during the study period (**Supplementary Table 1**). 47.4% of the full study population were male. Characteristics of the full study population are presented in **Table 1**.

**Table 1.**
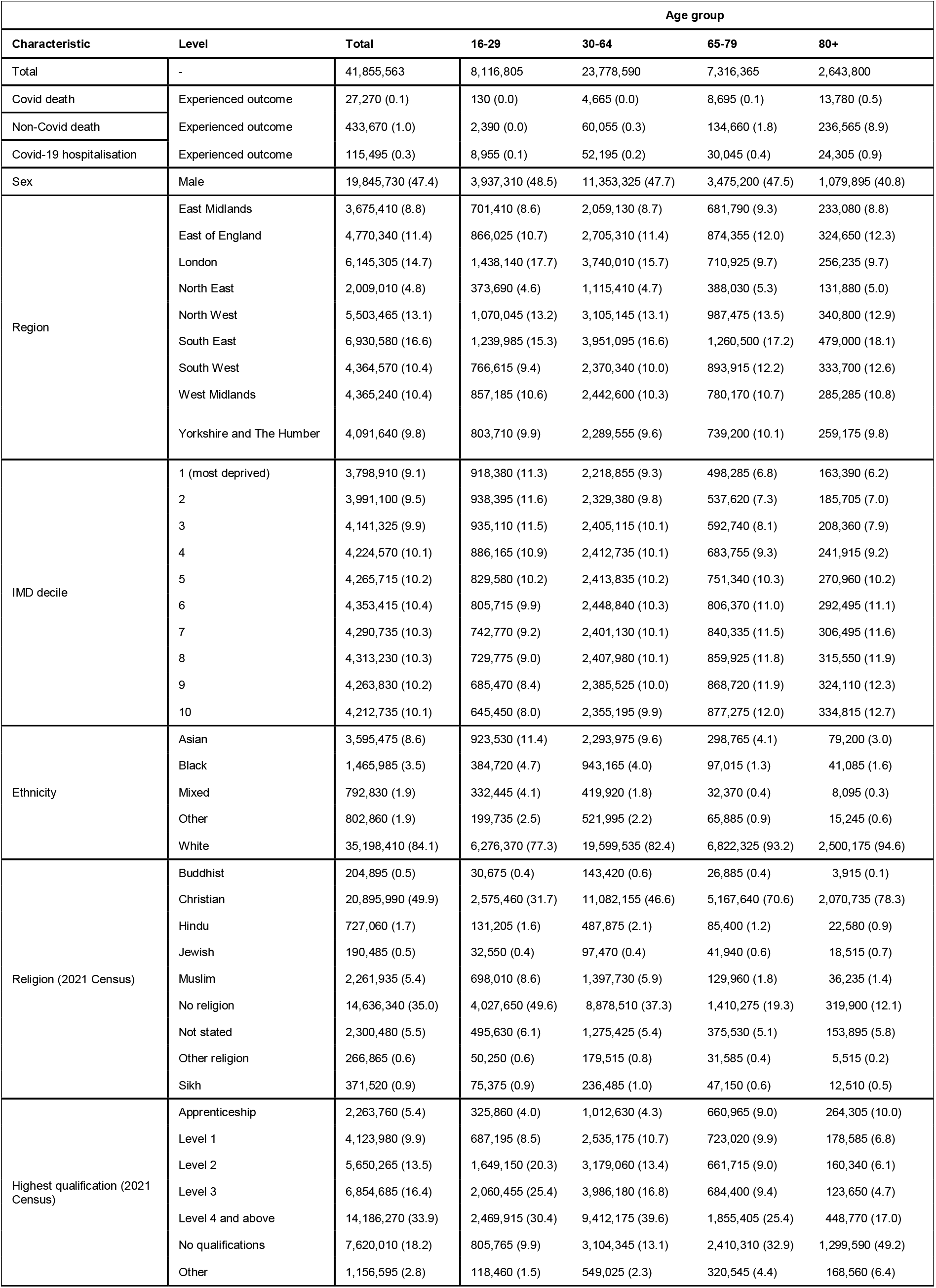
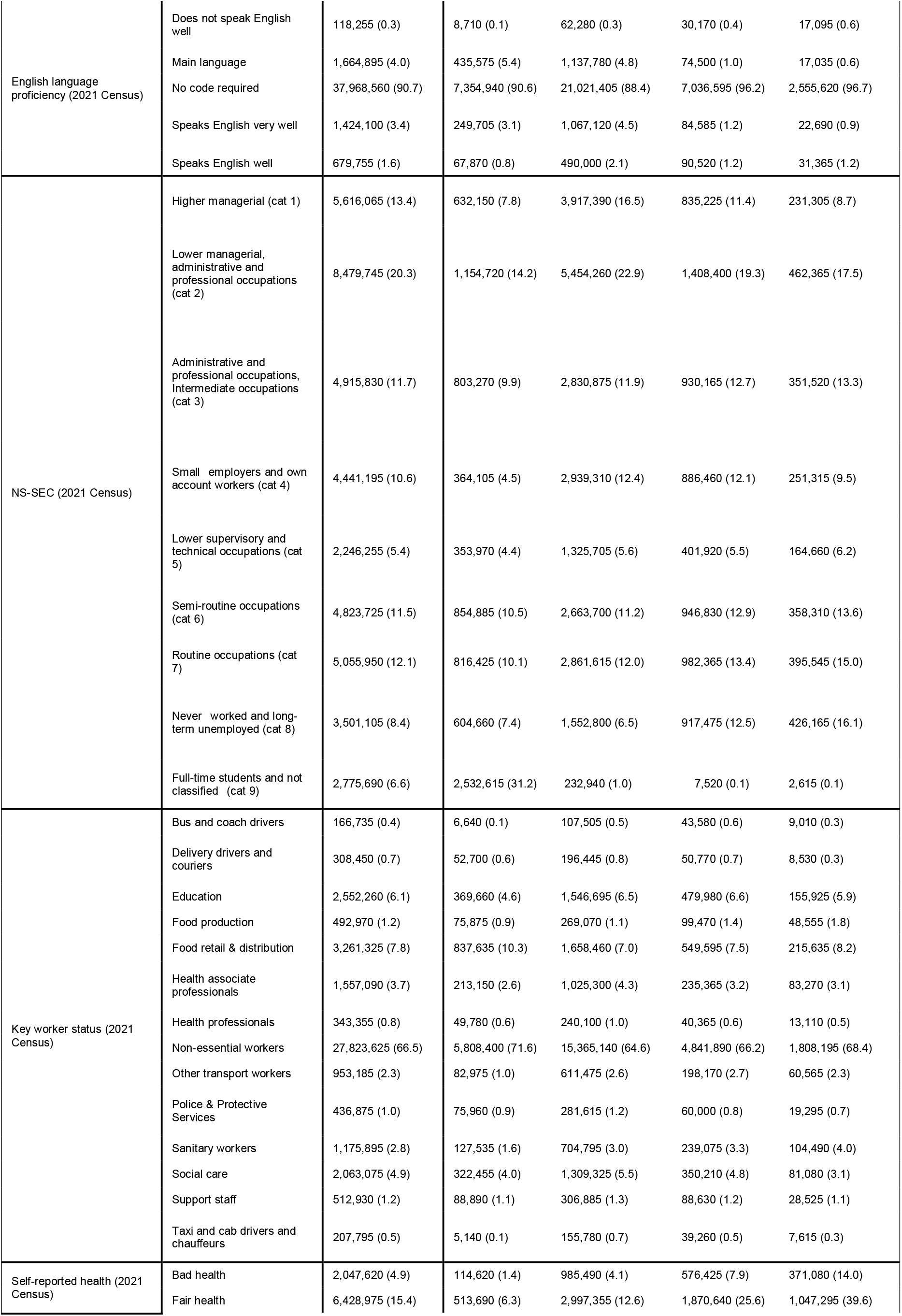
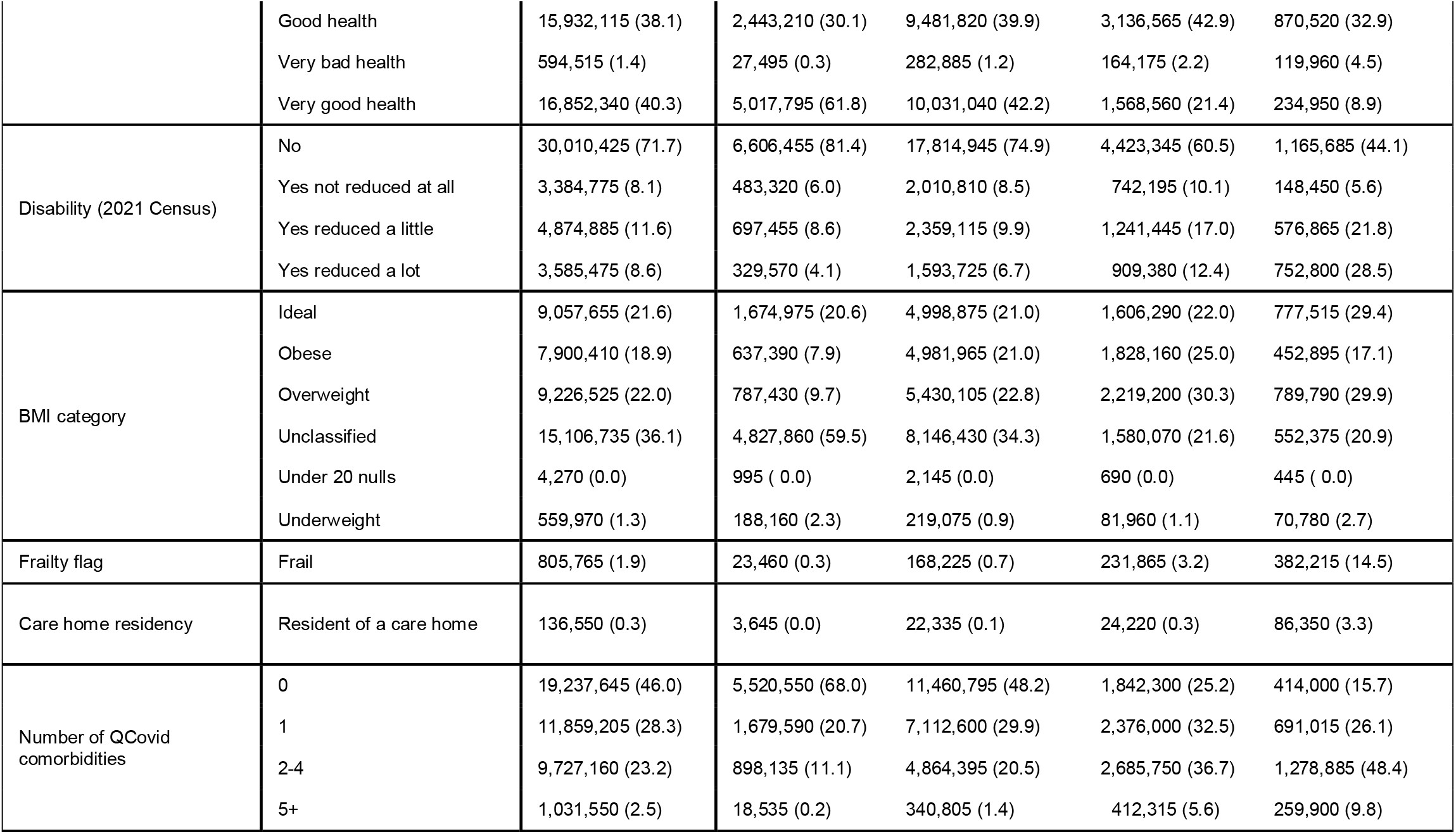
Characteristics of the full population, stratified by age-group.

#### Vaccine effectiveness against COVID-19 hospitalisation and death

Estimates of vaccine effectiveness (VE) against COVID-19 hospitalisation and death increased with the number of doses (**Figure 1, Table 2**). VE against hospitalisation after 3 doses was 77.6% (95% confidence interval; 77.3% to 78%), with some evidence of waning 3+ months after dose 3 (67.8% (67.0% to 68.5%)). VE increased 3-6 months and 6+ months after the second dose when compared to 21 days to 3 months after the second dose (**Table 2**). However, the estimate of VE was negative 3+ months after the first dose.

**Table 2.** Vaccine effectiveness against COVID-19 hospitalisation and death involving COVID-19.

| Vaccine status | Covid-19 Hospitalisation |  | Covid-19 death |  |
| --- | --- | --- | --- | --- |
|  | Hazard ratio | Vaccine effectiveness (%) | Hazard ratio | Vaccine effectiveness (%) |
| First dose 21 days to 3 months | 0.48 (0.47, 0.49) | 52.1 (51.3, 52.8) | 0.41 (0.36, 0.47) | 58.7 (52.7, 63.9) |
| First dose 3+ months | 2.00 (1.98, 2.02) | -100.36 (-102.45, -98.29) | 0.50 (0.46, 0.54) | 50.3 (46.3, 53.9) |
| Second dose 21 days to 3 months | 0.44 (0.44, 0.45) | 55.6 (55.2, 56.1) | 0.11 (0.11, 0.13) | 88.5 (87.5, 89.5) |
| Second dose 3-6 months | 0.33 (0.33, 0.34) | 66.7 (66.3, 67.1) | 0.16 0.15, 0.17) | 84.1 (83.4, 84.9) |
| Second dose 6+ months | 0.40 (0.40, 0.41) | 59.5 (58.98, 60.03) | 0.27 (0.26, 0.28) | 73.4 (72.28, 74.4) |
| Third dose 21 days to 3 months | 0.22 (0.22, 0.23) | 77.6 (77.3, 78.0) | 0.07 (0.06, 0.07) | 93.2 (92.9, 93.5) |
| Third dose 3+ months | 0.32 (0.32, 0.33) | 67.8 (67.0, 68.5) | 0.13 (0.12, 0.13) | 87.5 (86.8, 88.1) |

**Figure 1.**
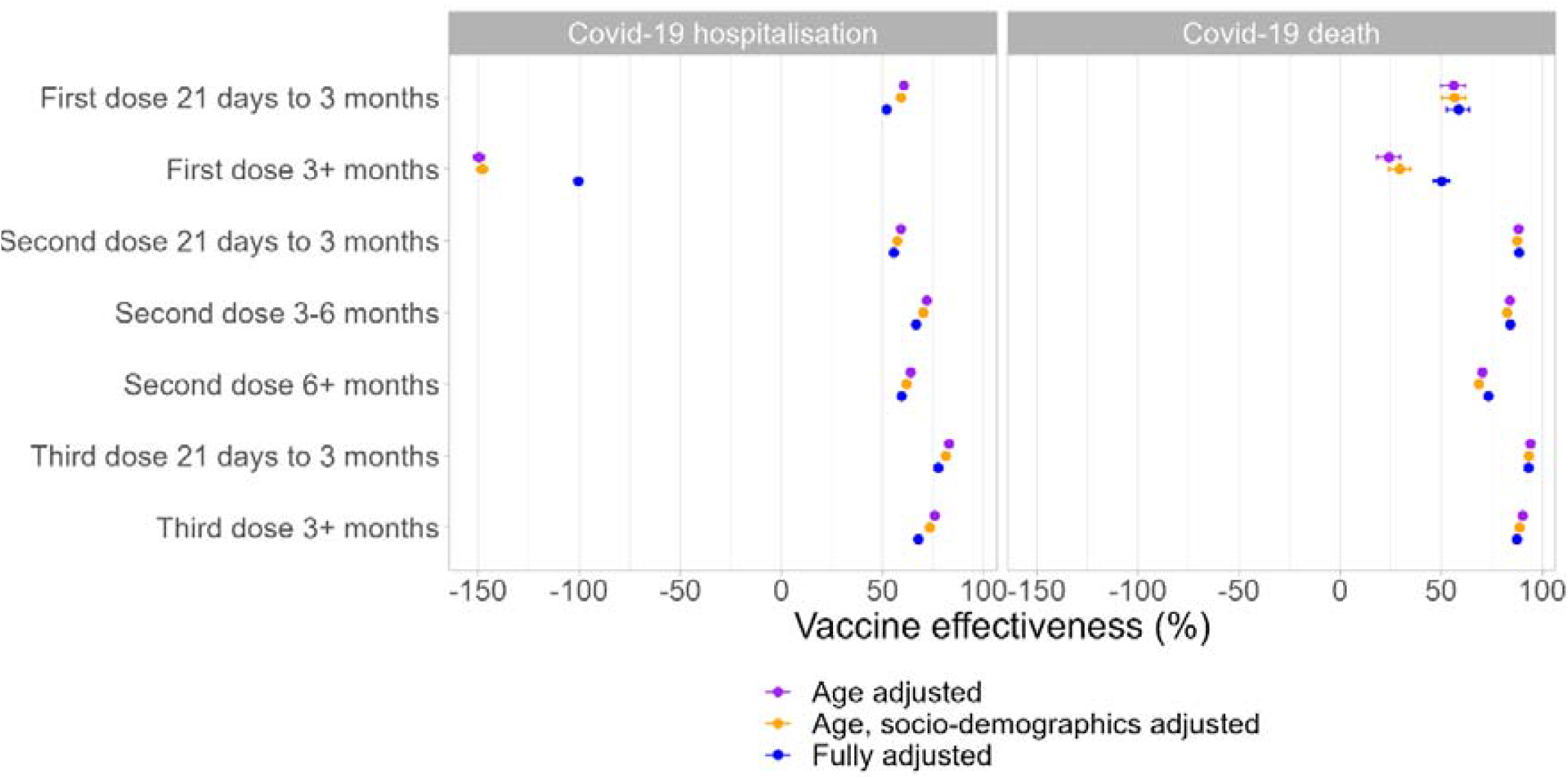
Vaccine effectiveness against COVID-19 hospitalisation and death involving COVID-19.

Again, VE against COVID-19 mortality increased with the number of doses (Table 2). VE against COVID-19 death after 3 doses was 93.2% (92.9% - 93.5%), with some evidence of waning after 3+ months, but effectiveness remained high.

#### Vaccine effectiveness against COVID-19 hospitalisation and death by age-group

VE against COVID-19 hospitalisation by age group followed similar trends to the non-age stratified VE among the older age groups (30-64, 65-79, 80+). The estimates of VE were negative for individuals in all age groups 3+ months after the first dose and in individuals aged 16-29, 3+ months after the third dose (**Table 3**). VE for the second and third doses was lowest for the 16-29 age group (**Table 3**). VE after the second dose was highest for the 30-64 (60.2% (59.5 to 60.8%)) and 65-79 (59.1% (58.3% to 59.9%)), age groups and VE for the third dose was highest for the 65-79 age group (87.5% (87.1% to 87.8%)). VE waned substantially for the 30-64 age group 3+ months after a third dose but remained high for the 65-79 and 80+ groups (**Table 3**).

**Table 3.** Vaccine effectiveness against COVID-19 hospitalisation and death involving COVID-19, stratified by age group.

| Age group | Vaccine status | Covid-19 Hospitalisation |  | Covid-19 death |  |
| --- | --- | --- | --- | --- | --- |
|  |  | Hazard ratio | Vaccine effectiveness (%) | Hazard ratio | Vaccine effectiveness (%) |
| 16-29 | First dose 21 days to 3 months | 0.56 (0.53, 0.59) | 44.2 (41.3, 47.0) | 0.44 (0.43, 0.45) | 56.1 (55.3, 57.0) |
|  | First dose 3+ months | 1.78 (1.69, 1.88) | -78.09 ( -87.65, -69.02) | 1.82 (1.79, 1.85) | -81.75 ( -84.94, -78.62) |
|  | Second dose 21 days to 3 months | 0.61 (0.59, 0.64) | 38.6 (35.7, 41.2) | 0.40 (0.39, 0.40) | 60.2 (59.5, 60.8) |
|  | Second dose 3-6 months | 0.59 (0.56, 0.62) | 40.9 (37.9, 43.7) | 0.30 (0.30, 0.31) | 69.6 (69.1, 70.2) |
|  | Second dose 6+ months | 0.90 (0.85, 0.96) | 9.5 (3.6, 15.0) | 0.45 (0.44, 0.45) | 55.5 (54.5, 56.5) |
|  | Third dose 21 days to 3 months | 0.82 (0.76, 0.89) | 18.0 (11.4, 24.1) | 0.29 (0.28, 0.30) | 70.7 (69.9, 71.5) |
|  | Third dose 3+ months | 2.33 (2.07, 2.62) | -132.77 (-161.90, -106.87) | 0.75 (0.72, 0.78) | 24.8 (21.6, 27.8) |
| 30-64 | First dose 21 days to 3 months | 0.44 (0.43, 0.45) | 56.1 (55.3, 57.0) | 0.19 (0.15, 0.23) | 81.5 (76.9, 85.2) |
|  | First dose 3+ months | 1.82 (1.79, 1.85) | -81.75 ( -84.94, -78.62) | 0.31 (0.26, 0.36) | 69.3 (63.9, 74.0) |
|  | Second dose 21 days to 3 months | 0.40 (0.39, 0.40) | 60.2 (59.5, 60.8) | 0.05 (0.05, 0.06) | 94.6 (93.6, 95.4) |
|  | Second dose 3-6 months | 0.30 (0.30, 0.31) | 69.6 (69.1, 70.2) | 0.10 (0.09, 0.11) | 89.9 (89.0, 90.8) |
|  | Second dose 6+ months | 0.45 (0.44, 0.45) | 55.5 (54.5, 56.5) | 0.20 (0.18, 0.22) | 79.7 (77.7, 81.5) |
|  | Third dose 21 days to 3 months | 0.29 (0.28, 0.30) | 70.7 (69.9, 71.5) | 0.06 (0.06, 0.07) | 93.7 (92.8, 94.5) |
|  | Third dose 3+ months | 0.75 (0.72, 0.78) | 24.8 (21.6, 27.8) | 0.15 (0.12, 0.18) | 85.0 (81.8, 87.6) |
| 65-79 | First dose 21 days to 3 months | 0.62 (0.59, 0.65) | 38.1 (35.2, 40.9) | 0.44 (0.34, 0.58) | 55.8 (42.3, 66.2) |
|  | First dose 3+ months | 1.75 (1.71, 1.78) | -74.70 ( -78.06, -71.41) | 0.45 (0.39, 0.52) | 54.8 (48.0, 60.7) |
|  | Second dose 21 days to 3 months | 0.41 (0.40, 0.42) | 59.1 (58.3, 59.9) | 0.09 (0.08, 0.11) | 90.8 (89.2, 92.3) |
|  | Second dose 3-6 months | 0.26 (0.25, 0.26) | 74.1 (73.6, 74.7) | 0.14 (0.12, 0.15) | 86.5 (85.3, 87.6) |
|  | Second dose 6+ months | 0.26 (0.26, 0.27) | 73.66 (73.00, 74.29) | 0.24 (0.23, 0.26) | 75.8 (73.9, 77.5) |
|  | Third dose 21 days to 3 months | 0.13 (0.12, 0.13) | 87.5 (87.1, 87.8) | 0.06 (0.05, 0.06) | 94.4 (93.8, 94.8) |
|  | Third dose 3+ months | 0.16 (0.15, 0.16) | 84.4 (83.7, 85.0) | 0.09 (0.08, 0.10) | 91.4 (90.5, 92.2) |
| 80+ | First dose 21 days to 3 months | 0.63 (0.59, 0.68) | 36.8 (32.3, 41.0) | 1.15 (0.92, 1.44) | -15.30 ( -44.23, 7.83) |
|  | First dose 3+ months | 1.74 (1.71, 1.78) | -74.10 ( -77.75, -70.53) | 0.74 (0.66, 0.83) | 25.7 (16.6, 33.9) |
|  | Second dose 21 days to 3 months | 0.46 (0.45, 0.47) | 54.0 (53.0, 55.0) | 0.22 (0.19, 0.26) | 77.9 (74.5, 81.0) |
|  | Second dose 3-6 months | 0.34 (0.33, 0.35) | 65.9 (65.2, 66.6) | 0.28 (0.26, 0.31) | 71.6 (69.2, 73.9) |
|  | Second dose 6+ months | 0.34 (0.33, 0.35) | 65.9 (65.1, 66.7) | 0.40 (0.38, 0.43) | 60.0 (57.3, 62.5) |
|  | Third dose 21 days to 3 months | 0.19 (0.19, 0.20) | 80.7 (80.1, 81.2) | 0.09 (0.08, 0.10) | 91.0 (90.3, 91.6) |
|  | Third dose 3+ months | 0.18 (0.18, 0.19) | 81.6 (80.9, 82.3) | 0.13 (0.12, 0.14) | 87.3 (86.3, 88.2) |

VE against COVID-19 mortality follows broadly similar trends to the non-age stratified VE among the older age groups (30-64, 65-79, 80+), however the VE after the first dose is not significantly different from zero, with larger uncertainty, for the 80+ age group (−15.3% (−44.2% to 7.8%)). VE for the second dose was high for both the 30-64 and 65 to 79 age groups, and lowest for the 80+ group (**Table 3**). VE after a third dose was highest for the 65 – 79 age group, but was high for all age groups, being over 90%, apart from the 16 – 29 age group, where it reached 70.7 % (69.9% to 71.5%). VE also remained high 3+ months after a third dose for all age groups, except the 16 – 29 age group, where substantially evidence of waning was present.

#### Vaccine effectiveness against COVID-19 hospitalisation and death by dominant variant-period

VE against COVID-19 hospitalisation during the Omicron-dominant period was lower for the first and third doses than for the pre-Omicron period, but higher for the second doses (**Table 4)**. However, there was a considerable decrease in VE 3-6 and 6+ months after the second dose in Omicron-dominant period where it decreased to only 29.5% (27.4% to 31.6%) after 6+ months. Conversely, third dose VE during the pre-omicron period was 80.2% (79.9% to 80.5%), compared with only 55.3% (54.1% to 56.5%) for the Omicron-dominant period. However, VE reduced to only 22.8% (9.7% to 34%) 3+ months after the third dose in the pre-Omicron period, whereas there was very little evidence of waning in the Omicron period. The negative VE observed in the main results for COVID-19 hospitalisation 3+ months after the first dose was only observed in the pre-omicron period (**Table 4**).

**Table 4.** Vaccine effectiveness against COVID-19 hospitalisation and death involving COVID-19, by variant period.

| Variant period | Vaccine status | Covid-19 Hospitalisation |  | Covid-19 death |  |
| --- | --- | --- | --- | --- | --- |
|  |  | Hazard ratio | Vaccine effectiveness (%) | Hazard ratio | Vaccine effectiveness (%) |
| Pre-Omicron | First dose 21 days to 3 months | 0.47 (0.46, 0.48) | 52.8 (52, 53.5) | 0.34 (0.3, 0.4) | 65.6 (59.9, 70.5) |
|  | First dose 3+ months | 1.99 (1.97, 2.01) | -99.4 (-102.5, -97.3) | 0.43 (0.4, 0.47) | 56.7 (52.7, 60.5) |
|  | Second dose 21 days to 3 months | 0.44 (0.43, 0.44) | 56.4 (56, 56.9) | 0.1 (0.09, 0.11) | 90.4 (89.4, 91.2) |
|  | Second dose 3-6 months | 0.32 (0.31, 0.32) | 68.4 (68, 68.7) | 0.13 (0.12, 0.13) | 87.2 (86.6, 87.8) |
|  | Second dose 6+ months | 0.35 (0.35, 0.35) | 65 (64.5, 65.4) | 0.17 (0.17, 0.18) | 82.5 (81.7, 83.4) |
|  | Third dose 21 days to 3 months | 0.2 (0.19, 0.2) | 80.2 (79.9, 80.5) | 0.03 (0.03, 0.04) | 96.6 (96.3, 96.9) |
|  | Third dose 3+ months | 0.77 (0.66, 0.9) | 22.8 (9.7, 34) | 0.11 (0.02, 0.78) | 89 (21.9, 98.5) |
| Omicron | First dose 21 days to 3 months | 0.71 (0.64, 0.8) | 28.6 (20.2, 36) | 0.77 (0.58, 1.02) | 22.9 (-2.5, 42) |
|  | First dose 3+ months | 0.68 (0.64, 0.73) | 31.6 (27.2, 35.8) | 0.64 (0.55, 0.74) | 36.2 (26.5, 44.7) |
|  | Second dose 21 days to 3 months | 0.24 (0.22, 0.26) | 76.1 (73.7, 78.2) | 0.27 (0.19, 0.37) | 73.2 (62.8, 80.6) |
|  | Second dose 3-6 months | 0.38 (0.36, 0.39) | 62.5 (60.6, 64.2) | 0.25 (0.2, 0.31) | 75.2 (69.5, 79.9) |
|  | Second dose 6+ months | 0.7 (0.68, 0.73) | 29.5 (27.4, 31.6) | 0.51 (0.48, 0.55) | 48.8 (45.3, 52.1) |
|  | Third dose 21 days to 3 months | 0.45 (0.44, 0.46) | 55.3 (54.1, 56.5) | 0.11 (0.11, 0.12) | 88.8 (88, 89.5) |
|  | Third dose 3+ months | 0.46 (0.44, 0.47) | 54.3 (52.9, 55.6) | 0.18 (0.17, 0.19) | 82.4 (81.2, 83.4) |

Further stratifying the results for COVID-19 hospitalisation by dominant variant period by age group (**Supplementary Figure 6**) showed that the negative VE in the pre-Omicron period 3+ months after the first dose was present across all age groups. However, the negative VE 3+ months after the third dose was driven by the 30-64 age group in the pre-Omicron period, with insufficient data in the 16-29 age group. Negative VE was also observed in the Omicron period for the 16-29 age groups, with low but positive VE for the 30-64 age group. Otherwise, the results for the dominant variant periods by age group were consistent with the results overall for the dominant variant periods.

VE against COVID-19 mortality during the Omicron-dominant period was lower for all doses compared with the pre-Omicron period, with very little protection against Omicron variants following a first dose. There was stronger evidence of waning for the second dose in the Omicron dominant period for 6+ months after the second dose (VE Omicron: second dose 6+ months 48.8% (45.3% to 52.1%).

#### Vaccine effectiveness against COVID-19 hospitalisation and death by vaccine vector

VE against COVID-19 hospitalisation is higher for mRNA vaccines than non-mRNA or unknown vaccines for all doses (**Table 5**). The mRNA vaccine vector types show evidence of waning 3+ months after the third dose, but neither vaccine vector type shows evidence of waning after the second dose.

**Table 5.** Vaccine effectiveness against COVID-19 hospitalisation and death involving COVID-19, by vaccine vector.

| Vaccine vector | Vaccine status | Covid-19 Hospitalisation |  | Covid-19 death |  |
| --- | --- | --- | --- | --- | --- |
|  |  | Hazard ratio | Vaccine effectiveness (%) | Hazard ratio | Vaccine effectiveness (%) |
| mRNA | First dose 21 days to 3 months | 0.33 (0.32, 0.34) | 66.7 (65.6, 67.8) | 0.40 (0.33, 0.48) | 60.5 (52.0, 67.4) |
|  | First dose 3+ months | 1.97 (1.94, 2.00) | -96.74 ( -99.63, -93.90) | 0.63 (0.56, 0.71) | 36.9 (28.9, 44.0) |
|  | Second dose 21 days to 3 months | 0.39 (0.38, 0.39) | 61.3 (60.8, 61.8) | 0.09 (0.07, 0.10) | 91.5 (90.0, 92.7) |
|  | Second dose 3-6 months | 0.30 (0.29, 0.30) | 70.1 (69.7, 70.5) | 0.12 (0.11, 0.13) | 88.2 (87.4, 88.9) |
|  | Second dose 6+ months | 0.38 (0.37, 0.38) | 62.4 (61.8, 63.0) | 0.24 (0.22, 0.25) | 76.5 (75.3, 77.6) |
|  | Third dose 21 days to 3 months | 0.22 (0.22, 0.23) | 77.6 (77.2, 77.9) | 0.07 (0.06, 0.07) | 93.2 (92.9, 93.5) |
|  | Third dose 3+ months | 0.32 (0.31, 0.33) | 67.9 (67.1, 68.6) | 0.13 (0.12, 0.13) | 87.4 (86.8, 88.1) |
| Not mRNA<br>or<br>unknown | First dose 21 days to 3 months | 0.54 (0.53, 0.55) | 46.0 (45.0, 46.9) | 0.42 (0.35, 0.50) | 58.4 (49.9, 65.4) |
|  | First dose 3+ months | 2.03 (2.01, 2.06) | -103.10 (-105.63, -100.60) | 0.44 (0.40, 0.48) | 56.1 (51.8, 60.1) |
|  | Second dose 21 days to 3 months | 0.48 (0.48, 0.49) | 51.6 (51.0, 52.2) | 0.12 (0.11, 0.14) | 87.7 (86.4, 88.9) |
|  | Second dose 3-6 months | 0.36 (0.36, 0.37) | 63.9 (63.4, 64.3) | 0.18 (0.17, 0.19) | 82.0 (81.0, 82.9) |
|  | Second dose 6+ months | 0.44 (0.43, 0.44) | 56.3 (55.6, 57.0) | 0.29 (0.28, 0.31) | 70.5 (69.2, 71.8) |
|  | Third dose 21 days to 3 months | 0.49 (0.43, 0.56) | 50.9 (44.0, 56.9) | 0.17 (0.13, 0.23) | 82.8 (76.6, 87.4) |
|  | Third dose 3+ months | 0.55 (0.42, 0.71) | 45.1 (28.6, 57.8) | 0.10 (0.05, 0.20) | 90.3 (79.7, 95.4) |

Similarly, VE against COVID-19 mortality was also higher for mRNA vaccines than non-mRNA or unknown vaccines for all doses. Both vaccine vector types show evidence of waning 3+ months after the second dose and mRNA vaccine vector types show evidence of waning 3+ months after the first dose

### Sensitivity tests

Vaccine effectiveness estimated against non-COVID-19 mortality resulted in VE that was significantly different from 0 for all doses and times since dose, with model adjustments reducing the difference and having a larger reducing effect than for VE against COVID-19 mortality (**Supplementary Figure 1**). VE against non-COVID-19 mortality by age group is negative 3+ months after the first dose overall and in all age groups and 21 days to 3 months after the first dose in age group 65-79.VE against non-COVID-19 mortality is also negative in the 16-29 age group 6+ months after the second dose (**Supplementary Figure 2**). VE against non-COVID-19 mortality by vaccine vector shows similar VE estimates by vaccine vector for the first dose and lower VE estimates for non mRNA and unknown vaccine vector types for the second and third doses. VE is negative 3+ months after the first dose of either vaccine type. Vaccine effectiveness 21 days to 3+ months after the third dose of a non mRNA or unknown vaccine is not significantly different from zero, however, the uncertainty for the third dose is high due to low numbers of third dose non mRNA vaccines administered.

Results for VE against COVID-19 hospitalisation omitting the time varying hospitalisation variable were consistent with the main analysis (**Supplementary Figure 3**).

Results for VE for all outcomes where the first 21 days after the first dose vaccination were excluded from the ‘unvaccinated’ category were consistent with the main results (**Supplementary Figure 4**).

## Discussion

To the best of our knowledge, this is the first study on COVID-19 vaccine effectiveness to use Census 2021 characteristics to attempt to adjust for residual confounding inherent with observational studies. Vaccine effectiveness (VE) against hospitalisation for COVID-19 was 52.1% (95% confidence interval 51.3% to 52.8%) for a first dose, 55.6% (55.2% to 56.1%) for a second dose and 77.6% (77.3% to 78.0%) for a third dose. Vaccine effectiveness against COVID-19 mortality was 58.7% (52.7% to 63.9%) for a first dose, 88.5% (87.5% to 89.5%) for a second dose and 93.2% (92.9% to 93.5%) for a third dose. Protection increased with number of doses, as expected from previous research. Our first dose VE estimates against hospitalisation were lower in general than other published estimates for the UK for all doses [6], [23]–[25]. Estimates of vaccine effectiveness against COVID-19 mortality were slightly lower than published estimates for the second and third doses [6], [24], [25], and much lower for the first dose [24]. This is likely to be due to the different time periods used, with follow up starting from 21 March 2021 rather than the start of the vaccination campaign on 8 December 2022. This would mean those in the oldest age groups who contributed person time to the post first dose intervals in this study received vaccination late and were likely a more vulnerable group who delayed vaccination due to poor health or were less able to visit vaccination sites, which initially were providing the Pfizer vaccine which was less transportable. This leads to low or negative estimates of VE as these vulnerabilities could not be adjusted for with the available covariates. The negative VE estimates 3+ months after the first dose are likely because this is after the time the second dose should have been given and likely includes many frail individuals who delayed or did not get a second dose due to ill health when it was scheduled. This negative VE at 3+ months post dose 1 for hospitalisation was not seen during the Omicron period, which is probably because by this time most of the person time in this period was a long time after 3 months and not close to when a second dose was scheduled and hence less subject to frailty bias at the time the second was scheduled, These biases post dose 1 also were seen to affect the non-COVID-19 death control outcome. Similarly, frailty bias could also affect the second dose results in older ages where people who are more frail have not received the booster vaccination when eligible, resulting in a lower VE against non-COVID-19 and COVID-19 mortality for 6+ months after the second dose. It is likely that the time varying-hospitalisation variable used to adjust for changes in health over time, which may cause people to delay vaccination, is insufficient to capture all time-varying confounding due to health.

Vaccine effectiveness against COVID-19 hospitalisation does not decrease with time since second dose overall, but does in the lower age groups, particularly the 16-29 age group. This decrease is observed in the omicron period, but not in the pre-omicron period. For the third dose, vaccine effectiveness against COVID-19 hospitalisation decreases after the third dose in younger age groups, but not in older age groups and the VE is negative 3+ months after the third dose in the 16-29 age group. Some of this decrease could be due to a healthy vaccinee effect, where people who are unwell are more likely to delay vaccination until better, which increases the VE for time periods sooner after vaccination compared to longer term. It could also be due to residual confounding by health status as the younger people who were vaccinated earlier, and who therefore make up a higher proportion of the later time since third dose vaccination statuses, were more likely to be clinically vulnerable. First booster vaccination for clinically extremely vulnerable people aged 16-49 began in September 2021, whereas booster vaccination for the non-clinically extremely vulnerable population aged 16-49 (who had not been eligible for earlier vaccination through their occupation or living with vulnerable people) began in late November 2021. Therefore the 3+ months after third dose category for 16-29 year olds will be skewed towards more at risk individuals as our study period ends on 20 March 2022. This issue is also seen when assessing the control outcome of non-COVID-19 mortality in 16-29 year olds. We see the VE in young people is particularly low in the pre-Omicron period for people aged 30-64 years (with insufficient data for 16-29 year olds) and then is particularly low for 16-29 year olds in the Omicron period. This provides further evidence for residual confounding by health status where there is prioritisation based on health status causing the negative VE as the larger impact moves to the group that will be most affected by prioritisation as the dominant variant period changes. Given the inconsistency of the decrease across the breakdowns, we cannot say there is strong evidence of waning protection against COVID-19 hospitalisation and confounding is likely to be impacting results. For VE against COVID-19 mortality, we do see consistent decreases across the various breakdowns for longer time periods after the second and third doses, where there is enough data to give a confident result. This could indicate waning of protection.

The sensitivity tests indicate that including the time-varying hospitalisation variable in the analysis of COVID-19 hospitalisation does not affect the estimates of VE. In addition, we also show in a sensitivity test that including the first 21 days after the first dose, when protection is rising, in the unvaccinated vaccination produces results consistent with the main analysis.

The results by time period show generally lower VE in the Omicron period for all doses against COVID-19 hospitalisation and mortality, except the second dose against COVID-19 hospitalisation. This appears inconsistent with research showing that the Omicron variant is less likely to cause severe outcomes. However, the first dose estimates are less reliable in the Omicron period and are for a small, unusual group who don’t get vaccinated when eligible and the third dose estimates are unreliable in the pre-omicron period as fewer people received the third dose or booster at this point.

The results by vaccine vector show generally lower results for non mRNA vaccines. However, this could be influenced by the different populations who received them. Of the third dose non-mRNA vaccines, very few of these will be booster vaccines. These are more likely to have been received by people who were given a third dose prior to booster vaccination due to being immunocompromised, so may represent a very different group to the general population.

Despite our inclusion of Census 2021 characteristics, health variables derived from ICD-10 codes and a time-varying flag for hospitalisation, estimates of VE against non-COVID-19 mortality are generally positive, indicating apparent VE which indicates the presence of residual confounding. For all doses, adjusting for common types of confounding improved estimates of VE, though only slightly in most cases, however we were unable to completely remove all forms of confounding within our analysis. Our adjustments had a greater impact on non-COVID-19 mortality, bringing the estimates closer to zero in most cases, indicating that confounding has more of an impact for non-COVID-19 mortality than COVID-19 outcomes. For all vaccination statuses, the VE against COVID-19 mortality is significantly higher than the VE against non-COVID-19 mortality.

In this study, we calculate the VE against hospitalisation and mortality based on the whole population rather than in those who are infected with COVID-19. This, combined with the use of death registrations to look at mortality rather than the definition of death within a certain timeframe of infection, means that VE against non-COVID-19 mortality can be calculated as a negative control outcome to assess the level of residual confounding that may be present. Though this study is not the first to look at COVID-19 vaccine effectiveness against non-COVID outcomes, we use this VE in order to explain the inherent issues with residual confounding in observational studies and caution against misinterpretation rather than report true effects.

This is the first study on COVID-19 vaccine effectiveness to use socio-economic and health variables from the 2021 census. This enables us to better adjust for confounding factors that other studies are not able to, and allows for more accurate estimates of vaccine effectiveness in a real-world setting.

The VE estimates presented here, produced with 2021 Census data, show increased protection with number of doses and a high level of protection against both COVID-19 hospitalisation and mortality for the third/booster dose, as would be expected from previous research. However, despite the various sources of health data used to adjust the models, the estimates for different breakdowns and for non-COVID-19 mortality expose residual confounding by health status, which should be considered in estimates of VE.

## Data Availability

All data produced in the present study are available upon reasonable request to the authors

