## Supplementary table for "Estimating the effectiveness of COVID-19 vaccination against COVID-19 hospitalisation and death: a cohort study based on the 2021 Census, England"

Supplementary Information

**Supplementary Table 1 | Sample flow**

| **Filters** | **Number of individuals** |
| --- | --- |
| Present in 2011 Census (not imputed), present in PDS and has a valid NHS number | 55,081,056 |
| Usual resident of UK and resident in England | 51,801,515 |
| Alive on 21 March 2021 | 51,797,278 |
| Age 16+ on 21 March 2021 | 41,950,323 |
| Non erroneous/ambiguous vaccination data | 41,855,563 |
| Sample of data | 583,840 |

**Supplementary Table 2: |Variables used in the analyses**

| **Variable** | **Coding** | **Source** |
| --- | --- | --- |
| **Outcomes** |  |  |
| Death involving COVID-19 | Confirmed or suspected COVID-19 (ICD -10 codes U07.1 and U07.2) as underlying or contributory cause of death on the death certificate | 2020 - 2022 ONS Death registrations |
| COVID-19 hospitalisation | An inpatient episode where either of the ICD10 codes, U07.1 (COVID-19, virus identified) or U07.2 (COVID-19, virus not identified) is the primary diagnosis. Where an individual has more than one COVID-19 hospitalisation, the earliest is used. | Hospital Episode Statistics |
| **Exposure** |  |  |
| Vaccination status | Time-updated covariate. Categories are:   - Unvaccinated (no vaccination to <21 days post first dose) - First dose (>= 21 days post second dose to earliest of < 91 days post first dose or < 21 days post second dose) - First dose 3+ months (>= 91 days post first dose to <21 days post second dose) - Second dose (>= 21 days post second dose to earliest of < 91 days post second dose or < 21 days post third dose) - Second dose 3-6 months (>= 91 days post second dose to earliest of < 182 days post second dose or < 21 days post third dose) - Second dose 6+ months (>= 182 days post second dose to < 21 days post third dose) - Third dose (>= 21 days post third dose to <91 days post third dose) - Third dose 3+ months (>= 91 days post third dose) | National Immunisation Management Service (NIMS) |
| **Confounding factors** | | |
| Age on 21 March 2021 | Natural spline, boundary knots at 10% and 90%, internal knots at 25%, 50% and 75% quantiles of the full dataset or the age specific datasets used for the age stratified results. | 2021 Census |
| Sex | Female, Male | 2021 Census |
| Self-reported ethnic group | Black,  Mixed, White, Asian, Other | 2021 Census |
| Religious affiliation | Christian, Buddhist, Hindu, Jewish, Muslim, Sikh, Other Religion, No religion, Religion Not Stated, No code required | 2021 Census |
| Region of residence in 2019 | Government Office region (E12000001-9) | Derived from 2021 Census OA |
| Index of Multiple Deprivation (IMD) | Binary variables representing deciles of deprivation | Derived from 2021 Census LSOA |
| Level of highest qualification | No qualification, Level 1 (1-4 GCSEs any grade or equivalent), Level 2 (5+ GCSEs grade A*-C or equivalent), Apprenticeship, Level 3 (2+ A Levels or equivalent), Level 4+ (Degree), Other, No code required | 2021 Census |
| English language proficiency | Can speak English very well, Can speak English well, Cannot speak English well, Cannot speak English, No code required | 2021 Census |
| National Statistics Socio-economic classification (NS-SEC) | The SOC 2020 classifications were grouped into the NS-SEC 8 analytic classes [26]:  Higher managerial (cat 1), Lower managerial, administrative and professional occupations (cat 2), Administrative and professional occupations, Intermediate occupations (cat 3), Small employers and own account workers (cat 4), Lower supervisory and technical occupations (cat 5), Semi-routine occupations (cat 6), Routine occupations (cat 7), Never worked and long-term unemployed (cat 8), Full-time students and not classified (cat 9) | 2021 Census |
| Key worker status | Based on SOC 2020 classification.  Taxi and cab drivers and chauffeurs, Support staff, Bus and coach drivers, Sanitary workers, Social care, Van drivers, Health associate professionals, Food retail & distribution, Other transport workers, Health professionals, Food production, Education, Police & Protective Services, Non-essential workers | 2021 Census |
| Care home residency | Binary variable for residency in a care home. 1 if the person is a resident in communal accommodation that is a local authority care home with nursing, a local authority care home without nursing, a care home with nursing or a care home without nursing. | 2021 Census |
| Long term health problem or disability | Day-to-day activities limited a lot, Day-to-day activities limited a little, Day-to-day activities not limited | 2021 Census |
| Self-reported general health | Very good health, Good health, Fair health, Bad health, Very bad health | 2021 Census |
| Body Mass Index (kg/m2) | Underweight (< 18.5), Ideal (18.5 – 25), Overweight (25 to 30), Obese (>= 30), Unclassified, Under 20 nulls | GDPPR V3 |
| Number of QCovid comorbidities | The QCOVID comorbidities included are:    Motor neurone disease, multiple sclerosis, myasthenia or Huntington’s Chorea; leukolaba; liver cirrhosis; asthma; prior fracture of hip, wrist, spine or humerous; Severe combined immunodeficiency; severe mental illness; epilepsy; cystic fibrosis or bronchiectasis alveolitis; heart failure; lung or oral cancer; pulmonary hypertension or fibrosis; Inflammatory bowel disease; prescription for prednisolone; HIV; dementia; diabetes type 1; diabetes type 2; chronic kidney disease; atrial fibrillation; schizophrenia; cancer of blood or bone marrow; Chronic obstructive pulmonary disease; rheumatoid arthritis or Systemic lupus erythematosus; cerebral palsy; Parkinson’s disease; learning disability or Down’s syndrome; thrombosis or pulmonary embolism; stroke or Transient ischaemic attack; immunosuppressants; congenital heart problem; coronary heart disease.    Grouping: 0, 1, 2-4, 5+. | GDPPR V3 |
| Frailty flag | Flag is 1 if individual experienced at least 1 hospital episode ending in the 5 years prior to 21 March 2021 where a primary or secondary diagnosis corresponding to frailty has been recorded. The ICD10 codes for frailty are as defined in the Hospital Frailty Risk Score [27]. | HES |
| Hospitalisation status | Binary, time-updated covariate. Status is 1 if currently hospitalised or if had a hospital spell that started less than 21 days prior to date. Hospitalisations with a primary diagnosis code of COVID-19 (U07.1 or U07.2), that are related to birth or pregnancy or that last less than 1 day are not included. | HES |
| Variant period | Time-varying covariate defined as ‘pre-omicron’ from start of study 21 March to 19 December 2021 and ‘omicron’ from 20 December 2021 to end of study 31 March 2022. | Created variable |
| Vaccine vector | Time-varying covariate for the vaccine vector type used for a particular vaccination status which changes at the same intervals as the time-varying covariate for vaccination status. Variable is a combination of the vaccination status variable and  either ‘none’ (if vaccination status is unvaccinated), ‘mRNA’ (vaccine is BNT162b2 (Pfizer-BioNTech) or Moderna) or ‘not mRNA or unknown’ (vaccine is ChAdOx1-S (Oxford/AstraZeneca), any other vaccine, or an unknown vaccine). | NIMS |

**Supplementary Table 3 | Key worker status SOC 2020 groupings**

| **Key worker status** | **SOC 2020 unit groups** | |
| --- | --- | --- |
| Health professionals | 2211 | Generalist medical practitioners |
|  | 2212 | Specialist medical practitioners |
|  | 2225 | Clinical psychologists |
|  | 2226 | Other psychologists |
|  | 2251 | Pharmacists |
| Health associate professionals | 2231 | Midwifery nurses |
|  | 2232 | Registered community nurses |
|  | 2233 | Registered specialist nurses |
|  | 2234 | Registered nurse practitioners |
|  | 2235 | Registered mental health nurses |
|  | 2236 | Registered children's nurses |
|  | 2237 | Other registered nursing professionals |
|  | - | - |
|  | 2254 | Medical radiographers |
|  | 2255 | Paramedics |
|  | 2256 | Podiatrists |
|  | 3219 | Health associate professionals n.e.c. |
|  | 3212 | Pharmaceutical technicians |
|  | 3213 | Medical and dental technicians |
|  | 1171 | Health services and public health managers and directors |
|  | 1231 | Health care practice managers |
|  | 2221 | Physiotherapists |
|  | 2259 | Other health professionals n.e.c. |
|  | 2222 | Occupational therapists |
|  | 2223 | Speech and language therapists |
|  | 2229 | Therapy professionals n.e.c. |
|  | 2252 | Optometrists |
|  | 2253 | Dental practitioners |
|  | 6133 | Dental nurses |
|  | 3240 | Veterinary nurses |
|  | 2240 | Veterinarians |
|  | 3211 | Dispensing opticians |
|  | 3214 | Complementary health associate professionals |
|  | 2224 | Psychotherapists and cognitive behaviour therapists |
|  | 2225 | Clinical psychologists |
|  | 2226 | Other psychologists |
|  | 7114 | Pharmacy and optical dispensing assistants |
| Support staff | 4211 | Medical secretaries |
|  | 6131 | Nursing auxiliaries and assistants |
|  | 6132 | Ambulance staff (excluding paramedics) |
|  | 7114 | Pharmacy and optical dispensing assistants |
|  | 9262 | Hospital porters |
| Social care | 1172 | Social services managers and directors |
|  | 1232 | Residential, day and domiciliary care managers and proprietors |
|  | 2461 | Social workers |
|  | 2463 | Clergy |
|  | 3221 | Youth and community workers |
|  | 2464 | Youth work professionals |
|  | 3223 | Housing officers |
|  | 2469 | Welfare professionals n.e.c. |
|  | 3229 | Welfare and housing associate professionals n.e.c. |
|  | 6134 | Houseparents and residential wardens |
|  | 6135 | Care workers and home carers |
|  | 6136 | Senior care workers |
|  | 6137 | Care escorts |
|  | 6232 | Caretakers |
|  | 6138 | Undertakers, mortuary and crematorium assistants |
| Education | 2311 | Higher education teaching professionals |
|  | 2312 | Further education teaching professionals |
|  | 2313 | Secondary education teaching professionals |
|  | 2314 | Primary education teaching professionals |
|  | 2315 | Nursery education teaching professionals |
|  | 2316 | Special and additional needs education teaching professionals |
|  | 2317 | Teachers of english as a foreign language |
|  | 2321 | Head teachers and principals |
|  | 2322 | Education managers |
|  | 2324 | Early education and childcare services managers |
|  | 2329 | Other educational professionals n.e.c |
|  | 1233 | Early education and childcare services proprietors |
|  | 2323 | Education advisers and school inspectors |
|  | 2319 | Teaching professionals n.e.c. |
|  | 6111 | Early education and childcare assistants |
|  | 3232 | Early education and childcare practitioners |
|  | 6112 | Teaching assistants |
|  | 3231 | Higher level teaching assistants |
|  | 6113 | Educational support assistants |
| Food and retail distribution | 7131 | Shopkeepers and owners - retail and wholesale |
|  | 5431 | Butchers |
|  | 5432 | Bakers and flour confectioners |
|  | 5433 | Fishmongers and poultry dressers |
|  | 7111 | Sales and retail assistants |
|  | 7112 | Retail cashiers and check-out operators |
|  | 7132 | Sales supervisors - retail and wholesale |
|  | 9241 | Shelf fillers |
|  | 9249 | Elementary sales occupations n.e.c. |
|  | 1242 | Managers in storage and warehousing |
|  | 1150 | Managers and directors in retail and wholesale |
| Food production | 8111 | Food, drink and tobacco process operatives |
|  | 1211 | Managers and proprietors in agriculture and horticulture |
|  | 1212 | Managers and proprietors in forestry, fishing and related services |
|  | 5111 | Farmers |
|  | 5112 | Horticultural trades |
|  | 5119 | Agricultural and fishing trades n.e.c. |
|  | 9111 | Farm workers |
|  | 9119 | Fishing and other elementary agriculture occupations n.e.c. |
| Taxi and cab drivers and chauffeurs | 8213 | Taxi and cab drivers and chauffeurs |
| Bus and coach drivers | 8212 | Bus and coach drivers |
| Van drivers | 8214 | Delivery drivers and couriers |
| Other transport workers | 1241 | Managers in transport and distribution |
|  | 8231 | Train and tram drivers |
|  | 6214 | Rail travel assistants |
|  | 8211 | Large goods vehicle drivers |
|  | 8234 | Rail transport operatives |
|  | 9211 | Postal workers, mail sorters and messengers |
|  | 6214 | Rail travel assistants |
|  | 8232 | Marine and waterways transport operatives |
|  | 8233 | Air transport operatives |
|  | 8239 | Other drivers and transport operatives n.e.c. |
|  | 8219 | Road transport drivers n.e.c. |
| Police and Protective Services | 1162 | Senior police officers |
|  | 1163 | Senior officers in fire, ambulance, prison and related services |
|  | 3311 | Non-commissioned officers and other ranks |
|  | 3312 | Police officers (sergeant and below) |
|  | 3313 | Fire service officers (watch manager and below) |
|  | 3314 | Prison service officers (below principal officer) |
|  | 3319 | Protective service associate professionals n.e.c. |
|  | 2462 | Probation officers |
|  | 6311 | Police community support officers |
| Sanitary workers | 9221 | Window cleaners |
|  | 9222 | Street cleaners |
|  | 9223 | Cleaners and domestics |
|  | 9225 | Refuse and salvage occupations |
|  | 9229 | Elementary cleaning occupations n.e.c. |
|  | 9224 | Launderers, dry cleaners and pressers |
| Non-essential workers |  | All other codes |

**Supplementary Figure 1 | Non-COVID-19 VE against death for the study population.** The model adjustments are age (as a natural spline), plus socio-demographic characteristics (sex, region, ethnicity, religion, IMD decile, NSSEC category, highest qualification, English language proficiency, key worker status), plus health-related characteristics (disability, self-reported health, care home residency, number of QCovid comorbidities (grouped), BMI category, frailty flag and hospitalisation within the last 21 days.

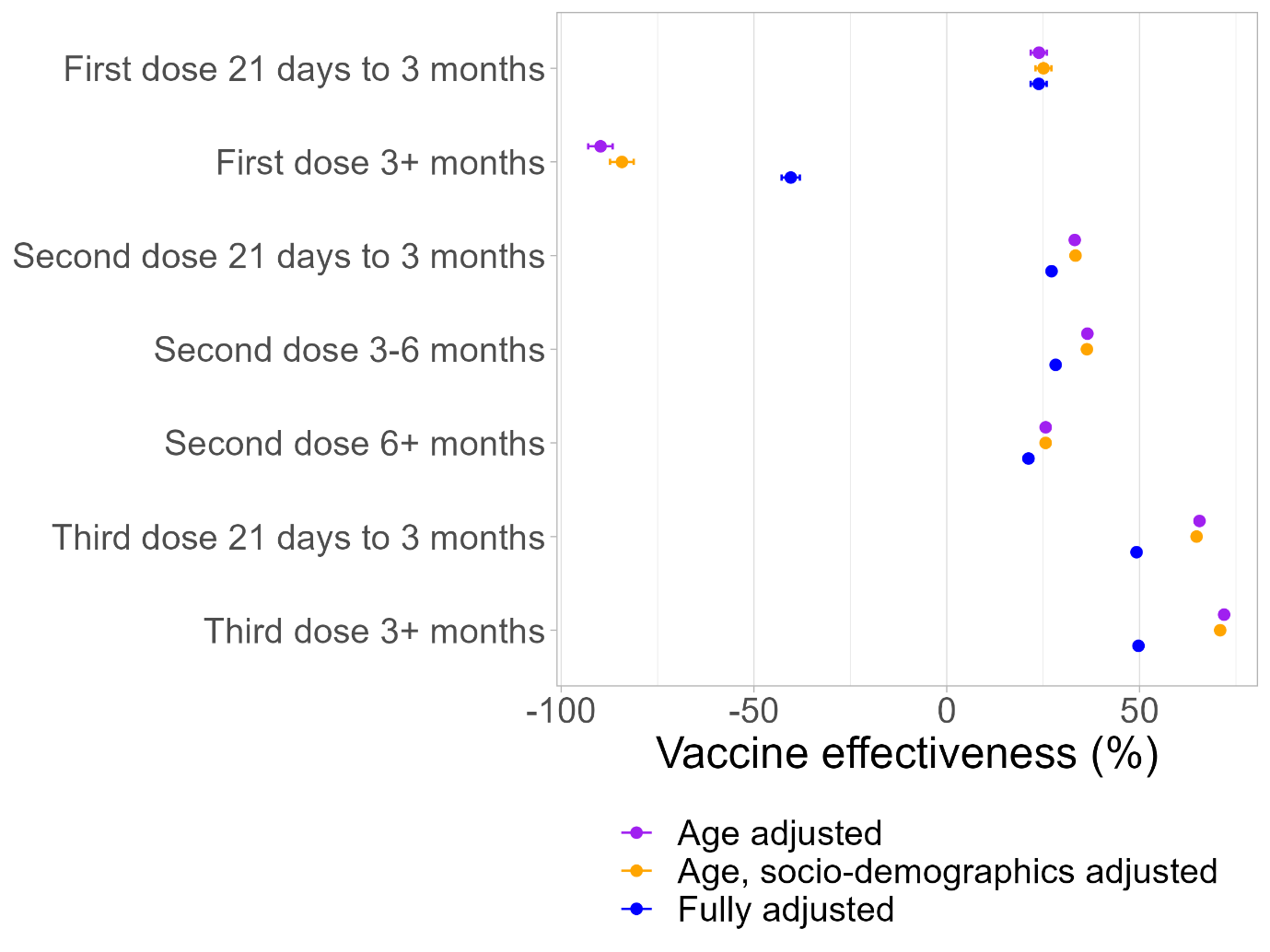

**Supplementary Figure 2 | Vaccine effectiveness against non-COVID-19 mortality, stratified by age group**. The model is fully adjusted. The covariates adjusted for are age (as a natural spline), sex, region, ethnicity, religion, IMD decile, NSSEC category, highest qualification, English language proficiency, key worker status, disability, self-reported health, care home residency, number of QCovid comorbidities (grouped), BMI category, frailty flag and hospitalisation within the last 21 days.

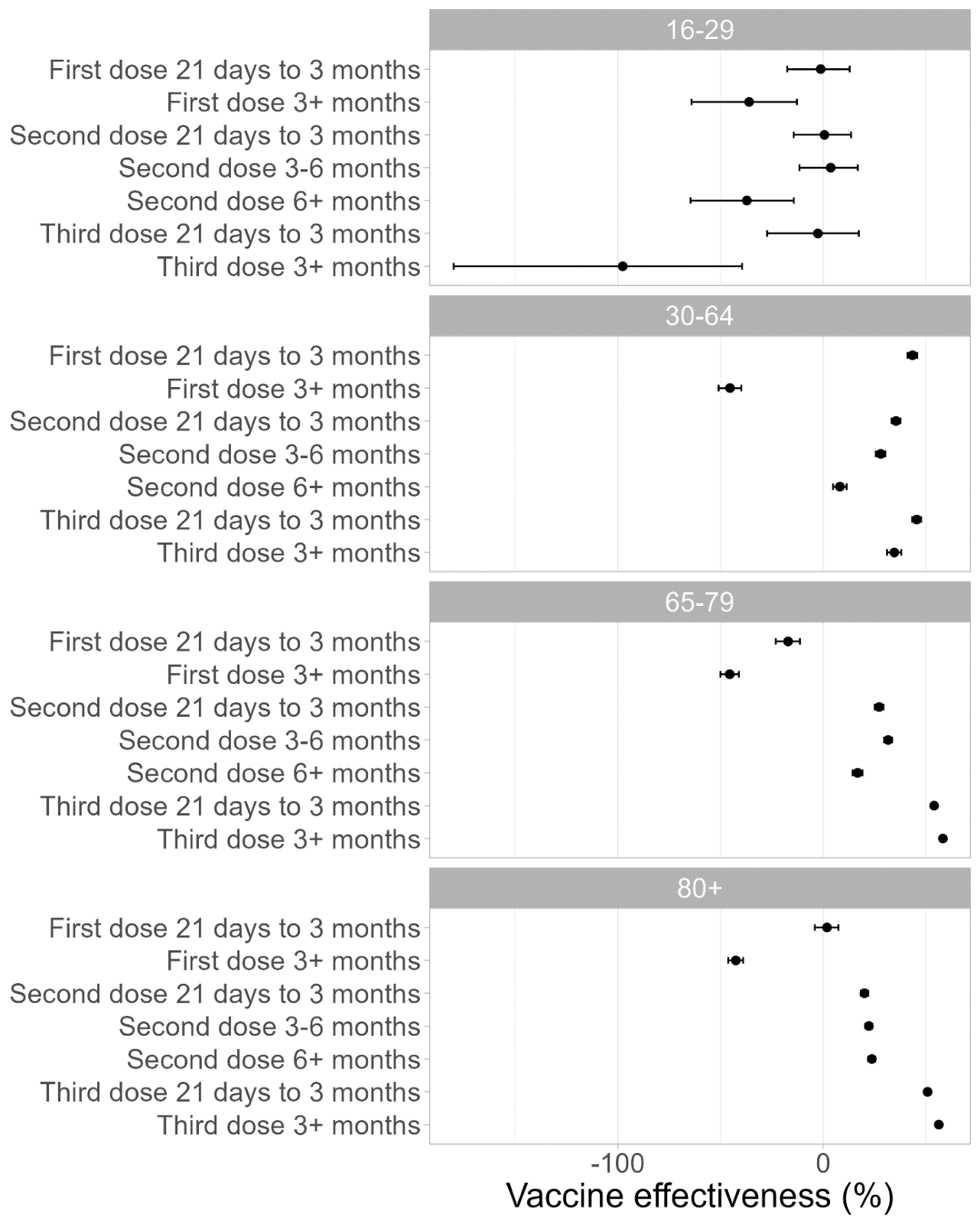

**Supplementary Figure 3 | Vaccine effectiveness against COVID-19 hospitalisation, omitting the time-varying variable for current hospitalisation or hospitalisation in the preceding 21 days**. All other covariates are adjusted for. These are age (as a natural spline), sex, region, ethnicity, religion, IMD decile, NSSEC category, highest qualification, English language proficiency, key worker status, disability, self-reported health, care home residency, number of QCovid comorbidities (grouped), BMI category and frailty flag.

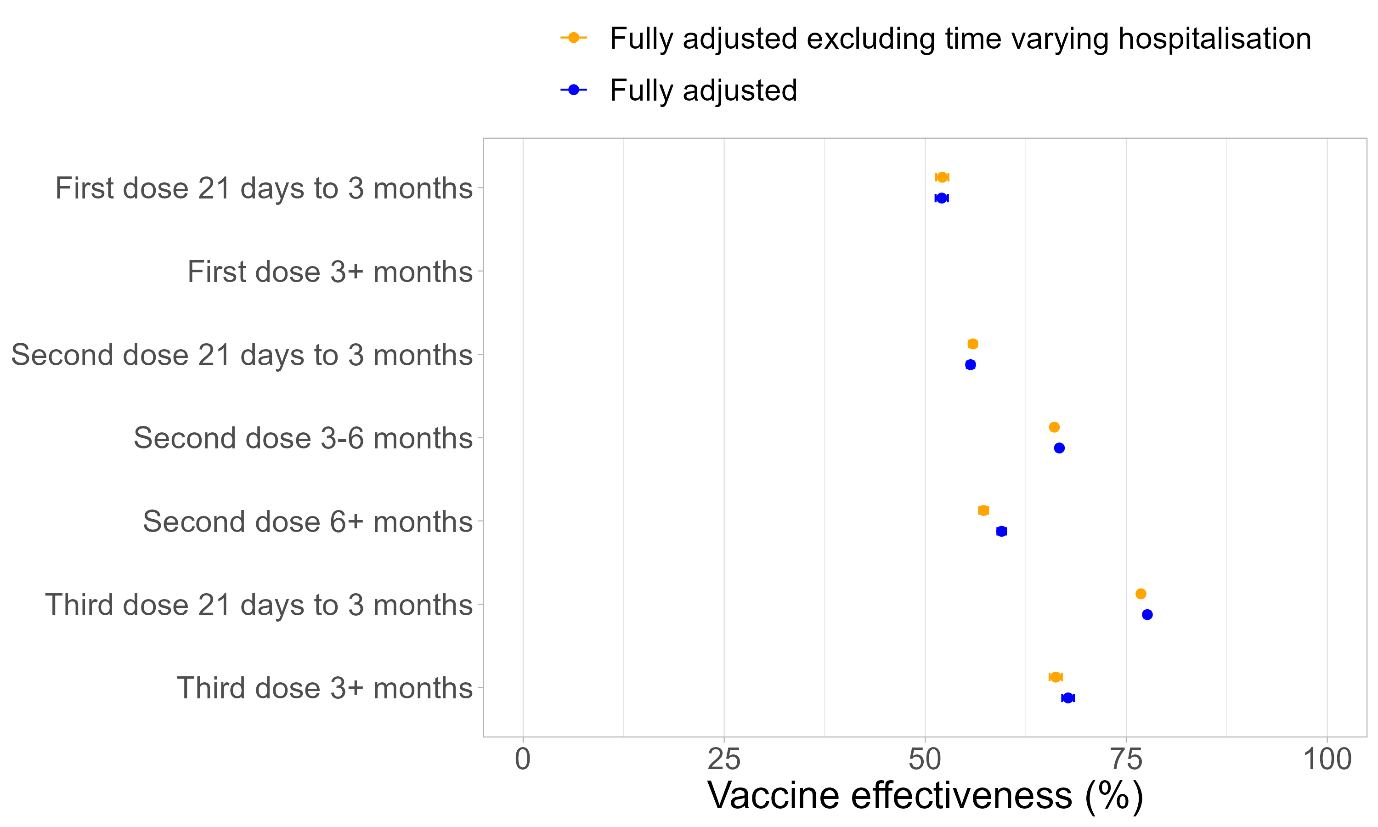

**Supplementary Figure 4 | Vaccine effectiveness against all outcomes, omitting the first 21 days after the first dose from the unvaccinated vaccination status**. The model was adjusted for age (as a natural spline), plus socio-demographic characteristics (sex, region, ethnicity, religion, IMD decile, NSSEC category, highest qualification, English language proficiency, key worker status), plus health-related characteristics (disability, self-reported health, care home residency, number of QCovid comorbidities (grouped), BMI category, frailty flag and hospitalisation within the last 21 days.

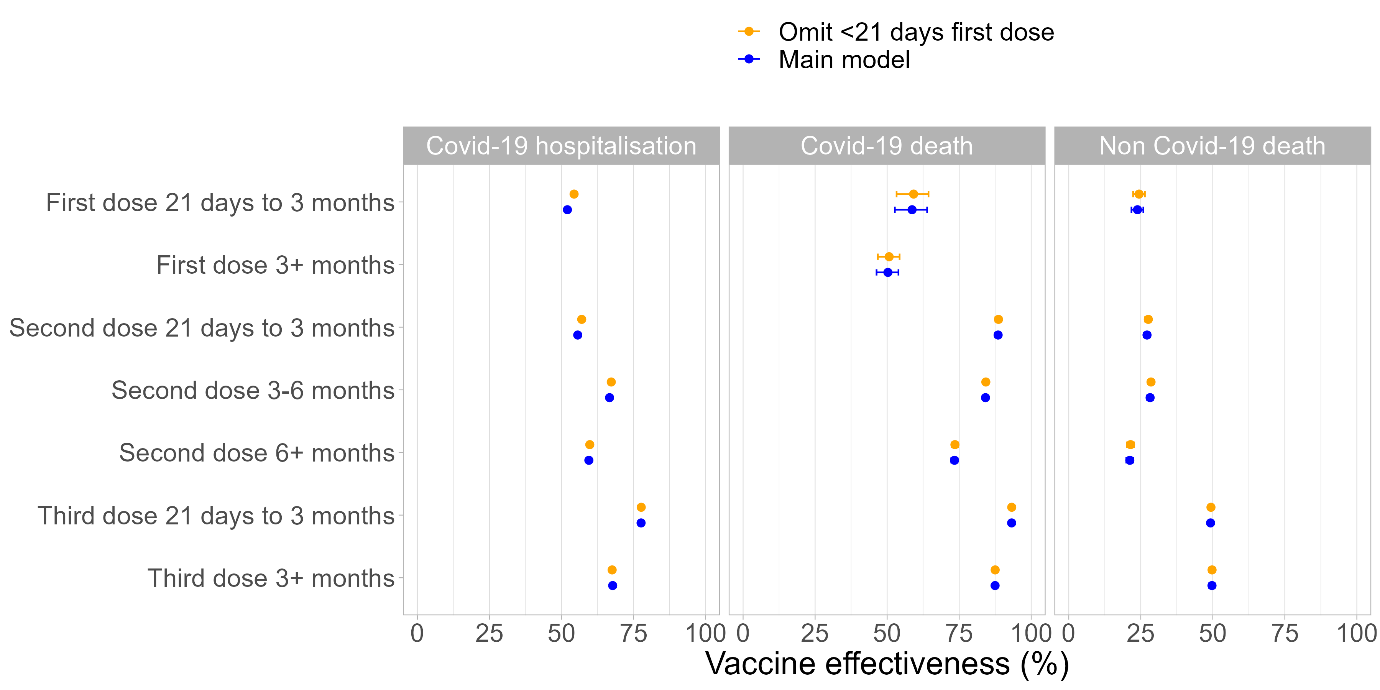

**Supplementary Figure 6 | Vaccine effectiveness against COVID-19 hospitalisation, by variant period, stratified by age group.** The model is adjusted for age (as a natural spline), sex, region, ethnicity, religion, IMD decile, NSSEC category, highest qualification, English language proficiency, key worker status, disability, self-reported health, care home residency, number of QCovid comorbidities (grouped), BMI category, frailty flag and hospitalisation within the last 21 days. Vaccination status is interacted with variant period.

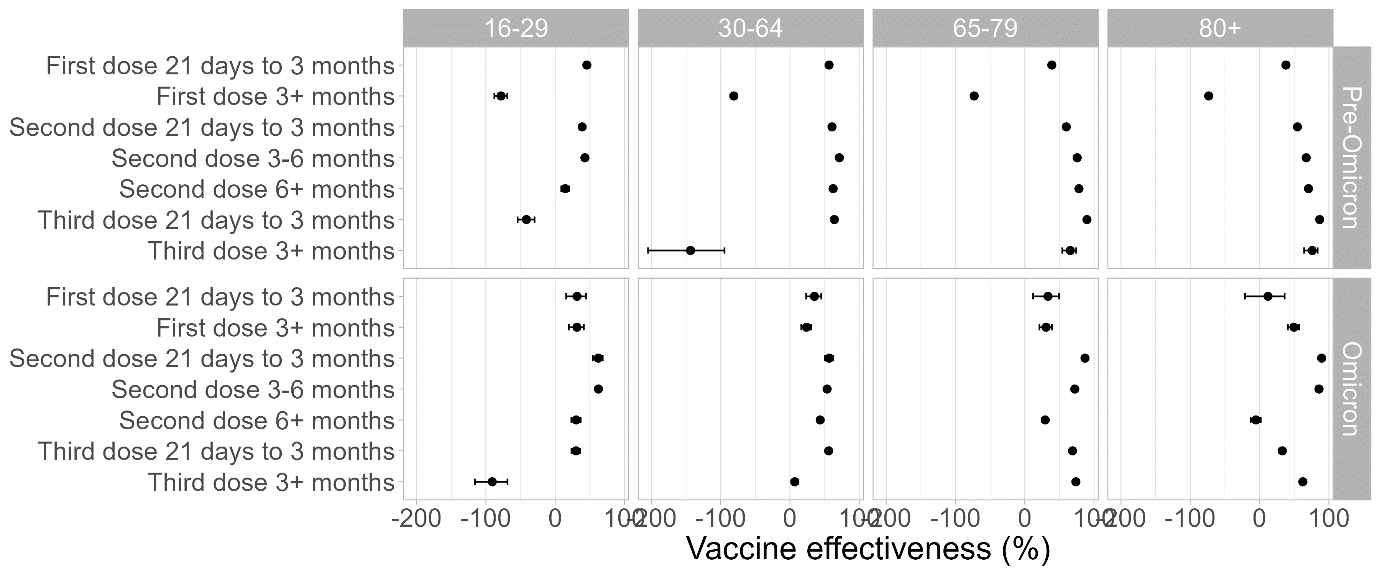
